# Prenatal Exposure to Early Life Adversity and Neonatal Brain Volumes at Birth

**DOI:** 10.1101/2021.12.20.21268125

**Authors:** Regina L. Triplett, Rachel E. Lean, Amisha Parikh, J. Philip Miller, Dimitrios Alexopoulos, Sydney Kaplan, Dominique Meyer, Chris Adamson, Tara A. Smyser, Cynthia E. Rogers, Deanna M. Barch, Barbara Warner, Joan L. Luby, Christopher D. Smyser

## Abstract

**Importance:** Exposure to early life adversity alters the structural development of key brain regions underlying neurodevelopmental impairments. The extent that prenatal exposure to life adversity alters structure at birth remains poorly understood.

**Objective:** To determine if prenatal exposure to maternal social advantage and psychosocial distress alters global and regional brain volumes and cortical folding in the first weeks of life.

**Design:** A prospective, longitudinal study of sociodemographically-diverse mothers recruited in the first trimester of pregnancy and their infants who underwent brain magnetic resonance imaging scan in the first weeks of life.

**Setting:** Mothers were recruited from local obstetric clinics from 2017-2020.

**Participants:** Of 399 mother-infant dyads prospectively recruited into the parent study, 280 healthy, term-born infants (47% female, mean postmenstrual age at scan 42 weeks) were eligible for inclusion.

**Exposures:** Maternal social advantage and psychosocial distress in pregnancy.

**Main Measures and Outcomes:** Two measures of latent constructs were created using Confirmatory Factor Analyses spanning Maternal Social Advantage (Income to Needs ratio, Area Deprivation Index, Healthy Eating Index, education level, insurance status) and Psychosocial Stress (Perceived Stress Scale, Edinburgh Postnatal Depression Scale, Everyday Discrimination Scale, Stress and Adversity Inventory). Neonatal cortical and subcortical gray matter, white matter, cerebellar, hippocampus, and amygdala volumes were generated using semi-automated age-specific segmentation pipelines.

**Results:** After covariate adjustment and multiple comparisons correction, greater social disadvantage (i.e., lower Advantage values) was associated with reduced cortical gray matter (*p*=.03), subcortical gray matter (*p*=.008), and white matter (*p*=.004) volumes and cortical folding (*p*=.001). Psychosocial Stress was not related to neonatal brain metrics. While social disadvantage was associated with smaller absolute volumes of the bilateral hippocampi and amygdalae, after correcting for total brain volume, there were no regional effects.

**Conclusions and Relevance:** Prenatal exposure to social disadvantage is associated with global reductions in brain volumes and cortical folding at birth. No regional specificity for the hippocampus or amygdala was detected. Results highlight that the deleterious effects of poverty begin *in utero* and are evident in the first weeks of life. These findings emphasize that preventative interventions to support fetal brain development should address socioeconomic hardships for expectant parents.

**KEY POINTS:** *Question:* Does prenatal exposure to maternal social disadvantage and psychosocial distress alter global and relative brain volumes at birth?

*Findings:* In this longitudinal, observational study of 280 mother-infant dyads, prenatal exposure to greater maternal social disadvantage, but not psychosocial distress, was associated with reduced white matter, cortical gray matter, and subcortical gray matter volumes and cortical folding at birth after accounting for maternal health and diet. There were no differential effects in the hippocampus or amygdala.

*Meaning:* Prenatal exposure to social disadvantage is associated with global reductions in brain volumes and folding in the first weeks of life.

## Introduction

Socioeconomic inequality is growing in the United States^1^ and places low-income, pregnant women at disproportionately greater risk of poor health outcomes^2^ and psychiatric disorders.^3,4^ Furthermore, low income is associated with higher levels of perceived stress during pregnancy.^5^ Thus, infants born to mothers experiencing social disadvantage are often exposed to multiple, overlapping socioenvironmental stressors.^6^ Early life adversity (ELA), including exposure to poverty, parental psychopathology, and stress, increases risk for adverse neurodevelopmental, socioemotional, and health outcomes from childhood and beyond.^7,8,9,10^

Human and animal studies suggest that altered brain development may be a key mechanism by which exposure to poverty and related psychosocial stressors increases the risk of poor outcomes.^8,11,12,13^ For example, poverty in early childhood is associated with reduced cortical gray and white matter, hippocampal, and amygdalae volumes at school-age.^14,15,16,17,18,19^ In turn, altered hippocampal volumes mediate the association of early adversity to behavioral problems in childhood.^20^ Despite clear and compelling negative impacts of social disadvantage on infant and child neurodevelopment,^7,8,9,10^ much less is known about its prenatal effects.

The prenatal period is a vulnerable stage of brain growth and development.^21,22^ The majority of neurogenesis and neuronal migration begins *in utero* with ongoing synaptogenesis, pruning, and myelination in the second and third trimesters.^23^ A small but growing literature demonstrates the deleterious and lasting consequences of prenatal exposure to maternal socioeconomic disadvantage and psychological stress on childhood outcomes, including cognitive delay, executive dysfunction, and higher rates of Conduct Disorder and Attention Deficit/Hyperactivity Disorder.^24,25,26^ However, few studies have explored the effect of prenatal ELA on brain outcomes at birth. The extant literature has largely conducted two parallel lines of research with most studies concentrating on maternal psychosocial stress, whereas few have examined prenatal exposure to social disadvantage.^27^ Further, the effects of related factors such as maternal health, diet, and substance use, have rarely been incorporated.

To date, studies investigating the effects of maternal mental health in pregnancy on neonatal brain volumes have focused on the hippocampus. Maternal depression and stress during pregnancy are associated with reduced hippocampal volumes and increased cortical folding.^28^ Other studies have shown that prenatal exposure to maternal depression and anxiety is associated with increased hippocampal volumes at birth and slower hippocampal growth.^29,30^ These findings highlight that subcortical structures may be particularly vulnerable to prenatal exposure to maternal mental health problems during a critical developmental window.

Far less is known regarding the role of social disadvantage during pregnancy on infant brain volumes at birth. One study found that lower socioeconomic status (indexed by income-to-needs ratio and education) was associated with smaller cortical and subcortical gray matter volumes at age one month,^31^ whereas Knickmeyer et al. found that maternal education but not income was positively associated with total white and gray matter volumes at birth.^32^ Knickmeyer et al. also found that independent of maternal education, maternal smoking in pregnancy and positive maternal psychiatric history explained variability in neonatal brain volumes.^32^ With the exception of this investigation, prior prenatal ELA studies have not considered the influence of maternal health, diet, and substance use, which may also impact brain structural growth and development among socially disadvantaged infants.^27,32,33,34,35,36^

In this study, we investigated the extent to which measures assessing latent constructs of maternal social advantage (including the contribution of diet) and psychosocial stress related to neonatal brain volumes and folding at birth. We included maternal health and substance use (tobacco and marijuana) as covariates. We analyzed global measures of cortical and subcortical gray matter, white matter, and cerebellar volume along with two structures of interest, the amygdala and hippocampus. We hypothesized that varying forms of prenatal ELA (i.e., reduced maternal social advantage and higher psychosocial stress) would each be associated with lower neonatal brain volumes, including regionally-specific susceptibility in the hippocampus and amygdala, and reduced cortical folding. Disentangling the independent roles of social disadvantage and maternal psychosocial distress *in utero* is vital to design targeted and effective preventative interventions that support fetal neurodevelopment.^11,21^

## Methods

### Study Design and Population

In this longitudinal, observational, multi-wave, multi-method NIMH-funded collaborative study, a cohort of pregnant women who participated in a large-scale study of preterm birth within the March of Dimes Prematurity Research Center at Washington University in St. Louis were recruited from 2017-2020. Women from the parent study were invited to participate in this investigation (see Luby, et al. for cohort details)^37^ with the following exclusion criteria: multiple gestations, diagnosed infections known to cause congenital disease (e.g., toxoplasmosis), and/or alcohol or drug use (excluding tobacco and marijuana). Participating mothers completed assessments during each trimester of pregnancy and at delivery. Medical data were collected from participant questionnaires and chart review. Neonatal brain MRI scans were performed in the first weeks of life. Exclusion criteria for the current analyses included premature birth (<37 weeks gestational age), Neonatal Intensive Care Unit admission >7 days, birthweight <2000g, or evidence of brain injury on MRI.

The study was approved by the Washington University Human Research Protection Office. Informed consent was obtained for each participant and subsequent parental informed consent was obtained for each infant prior to participation.

### Measures

#### Demographics, Social Advantage, and Psychosocial Stress

Maternal age, health insurance status (public, private, uninsured), highest educational level, address, and household composition were obtained from all participants each trimester during pregnancy. Participants also reported their self-identified racial/ethnic background.

As described in Luby et al., confirmatory factor analysis was used to derive two composite measures of Maternal Social Advantage and Maternal Psychological Stress.^37^ The following datapoints were used to estimate Social Advantage: household Income to Needs Ratio (I/R)^38^ in each trimester, national Area Deprivation Index (ADI) percentiles,^39^ Healthy Eating Index (HEI),^40^ highest level of educational attainment, and health insurance status. Race was highly correlated with Social Advantage, offering no additional improvement to the model after other variables (including racial discrimination) were accounted for and, thus, it was not included in the latent Advantage composite. To estimate Psychosocial Stress, four measures were used: Perceived Stress Scale (PSS) in each trimester,^41^ Edinburgh Postnatal Depression Scale (EPDS) in each trimester,^42^ Stress and Adversity Inventory (STRAIN, lifetime count and severity) at time of neonatal scan (*n*=190) or at follow-up at one or two years (*n*=80),^43^ and Everyday Discrimination Scale (EDS) at time of neonatal scan.^44^

#### Maternal Medical Co-Morbidities

A Maternal Medical Risk (MMR) score was calculated for each participant using questionnaire data and chart review.^45^ This validated index^46^ is a sum of weighted maternal co-morbidities including advanced age, cardiac disease, and pre-eclampsia with higher scores predicting increased risk of severe maternal morbidity (acute end-organ injury) or mortality. Maternal pre-pregnancy BMI is not included in the index and was therefore independently evaluated as a co-variate of interest.

#### Magnetic Resonance Imaging (MRI) Scanning

All MRI scans occurred within the first weeks of life without sedation during natural sleep. MRI data were collected using a Siemens Prisma 3T scanner and a 64-channel Siemens head coil. T1- and T2-weighted and spin echo fieldmap data were acquired with the following sequence parameters, T1: repetition time (TR)=2400ms, echo time (TE)=2.22ms, voxel size=0.8×0.8×0.8 mm^3^; T2: TR=3200/4500ms, TE=563ms, tissue T2=160ms, voxel size=0.8×0.8×0.8 mm^3^, and spin echo: TR=8000ms, TE=66ms, voxel size=2×2×2 mm^3^; 2 mm isotropic, multiband factor (MB)=1. Infants (n=11) without high-quality (i.e., low motion) structural data as determined by an experienced rater (DA) were excluded.

#### MRI Preprocessing and Brain Volumetric Measures

T1 and T2 images were corrected for gradient nonlinearity and readout distortions using FSL tools^47^ then denoised using ANTs.^48^ The Melbourne Children’s Regional Infant Brain atlas Surface (M-CRIB-S) segmentation and surface extraction toolkit was used to generate segmentations and surface-based cortical parcellations from preprocessed T2 images.^49,50^ The M-CRIB-S toolkit included N4 bias field correction and brain extraction, as well as automatic segmentation into white and gray matter, cerebellum, brainstem, and subcortical gray matter subdivisions corresponding to FreeSurfer-like labeling. Curvature-based spherical registration and mapping, alignment, and averaging were performed allowing for spatial normalization within the cohort and to the M-CRIB atlas. All segmentations and surfaces were inspected and manually corrected for accuracy by two experienced raters (DA and DM).

Brain volumes of interest included total cortical and subcortical gray matter, white matter, and cerebellum, in addition to right and left hippocampi and amygdalae. Total volumes for all structures were analyzed, as were standardized regional volumes for the hippocampi and amygdalae generated by dividing by total brain volume (TBV). Cortical folding was measured using the total surface area of the cortex for both hemispheres.

#### Statistical Analysis

Analyses were performed using SPSS version 27 (IBM Corporation, New York). Potential co-variates were explored using Pearson’s correlation and t-tests. Co-variates that demonstrated significant associations with brain volumes of interest included maternal tobacco use, infant sex, birthweight, and postmenstrual age (PMA) at MRI scan (Table S2). These variables and the factor scores for Social Advantage and Psychosocial Stress were included as independent variables in hierarchical regression analyses, each with regional brain volumes/cortical folding as the dependent variable. For each volume of interest, the first step accounted for maternal tobacco use, infant sex, birthweight, and PMA at MRI scan. The Social Advantage and Psychosocial Stress factors were added simultaneously in the second step of the model to determine the unique, independent proportion of variance explained in brain volume and folding outcomes over and above covariate factors. Regression models were checked for linearity, homoscedasticity, absence of multicollinearity, and the residuals approximated a normal distribution. Results for primary outcomes (total volumes, relative hippocampal and amygdalae volumes, and cortical folding) were corrected for multiple comparisons using a Benjamini-Hochberg False Discovery Rate procedure.^51^

## Results

The study recruited *n*=395 women during pregnancy (*n*=268 eligible subjects declined participation) and their *n*=399 singleton offspring (*n*=4 mothers had 2 singleton births during the recruitment period). After applying exclusion and data quality criteria, 280 mother-infant dyads were included in current analysis (see Figure S1 for exclusion reasons).

### Infant Clinical Characteristics

Infants were born at 39 weeks gestation on average (range 37–41) with 53% males (Table 1). At the time of MRI scan, infants were at an average of 42 weeks PMA (range 38–45). Male infants had larger birthweights than females (*p*=.03, Table S1). There were no additional sex differences for scan PMA, Social Advantage, and Psychosocial Stress (Table S1).

**Table 1.**
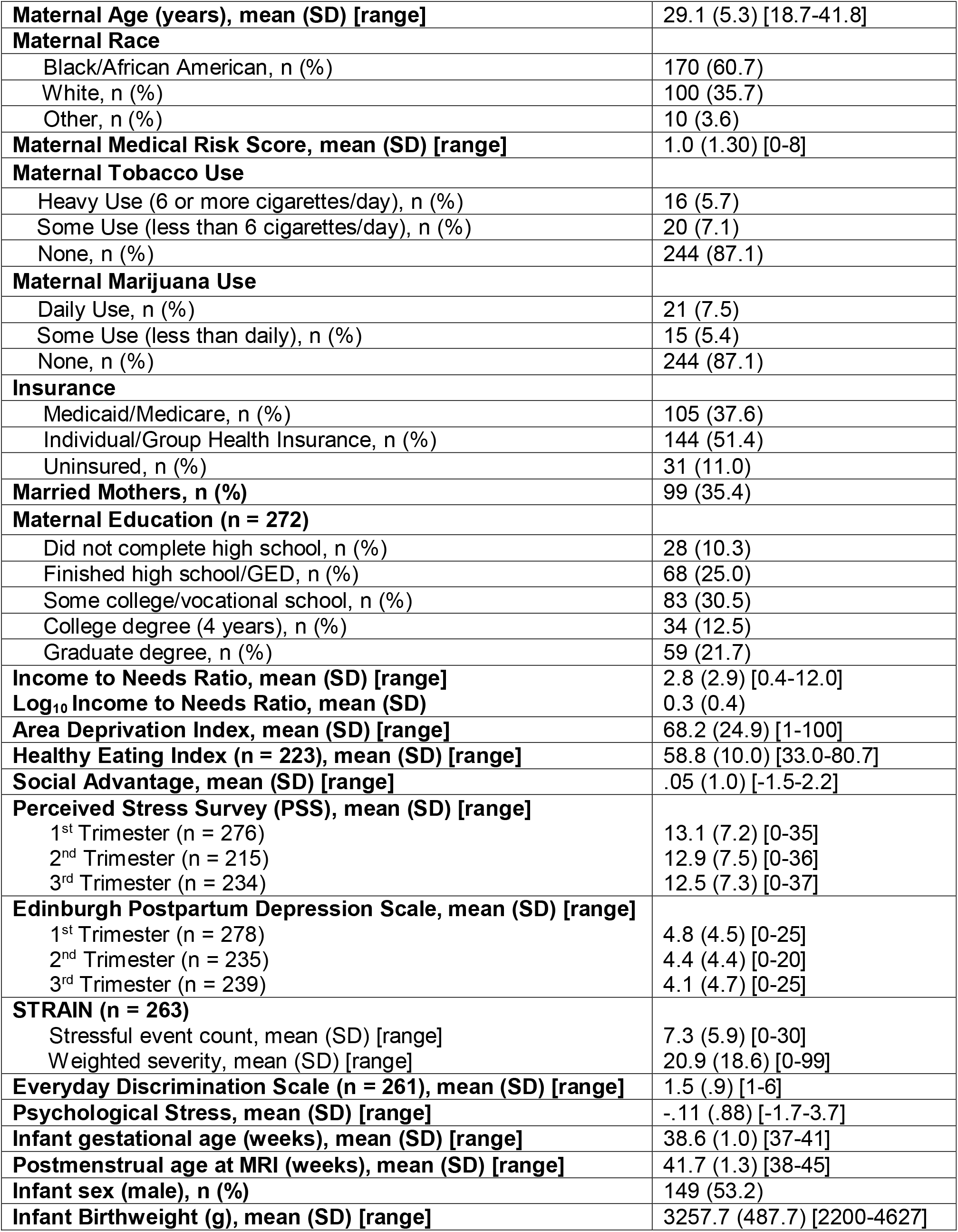
Social Background and Infant Clinical Characteristics of the Sample (*n*=280)

### Prenatal Life Adversity

Table 1 provides a summary of the prenatal life adversity characteristics of the sample, including the latent constructs of maternal Social Advantage and Psychological Stress. Higher Social Advantage was associated with lower maternal Psychological Stress (*r*=-0.42, *p*<.001).

### MRI Volumes

#### Brain Volumes

Table 2 provides a summary of the second, final step of the hierarchical regression results, with full results in Table S3. In Step 1, the following co-variates were related to smaller cortical and subcortical gray matter, white matter, and cerebellar volumes: female sex, lower birthweight, and younger PMA at scan (Table S3). Tobacco use related to reduced subcortical gray and white matter (Table S3). In Step 2, greater social disadvantage (i.e., lower Social Advantage values) related to reduced volumes across all tissue types and accounted for an additional 1.6-7% of the variance (Table 2, Figure 1) except for the cerebellum which did not survive correction for multiple comparisons (Table S3). The contribution of Psychosocial Stress was not significant (Table 2). A similar pattern of results was found for TBV (Table S4).

**Table 2.**
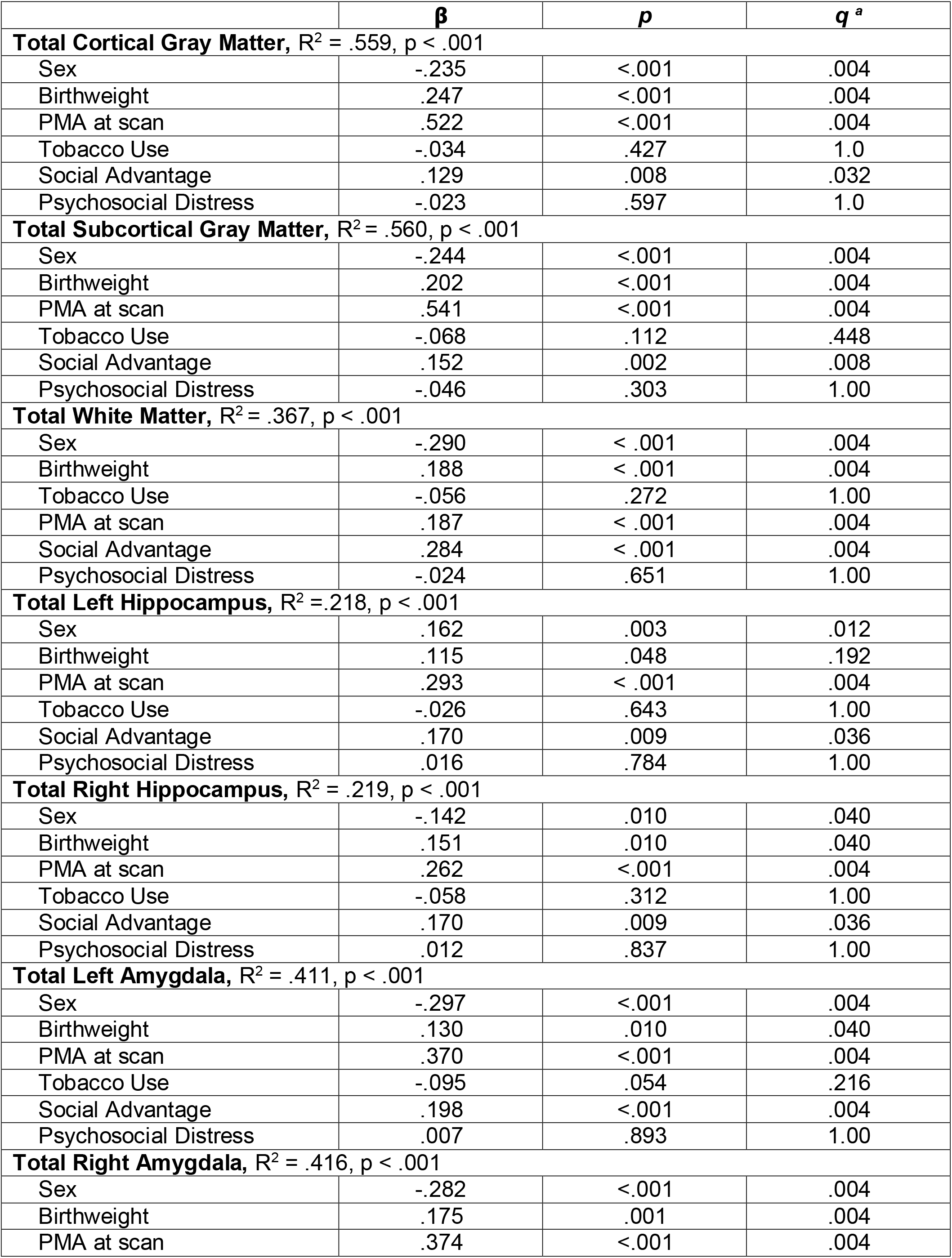
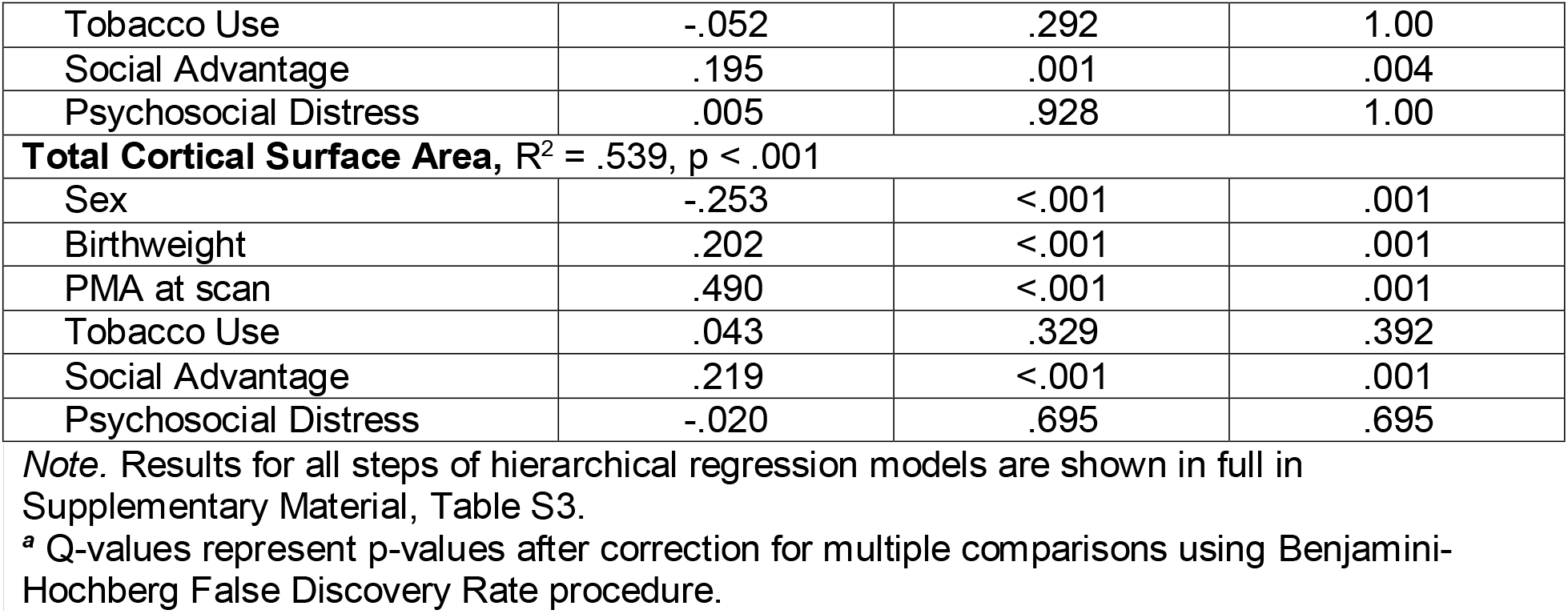
Summary of Final Step in Hierarchical Regression Models Linking Maternal Social Advantage and Psychosocial Stress with Structural MRI Measures at Birth (*n*=280)

| | $\beta$ | $p$ | $q^a$ |
| --- | --- | --- | --- |
| **Total Cortical Gray Matter, $R^2 = .559$, $p < .001$** |  |  |  |
| Sex | -.235 | <.001 | .004 |
| Birthweight | .247 | <.001 | .004 |
| PMA at scan | .522 | <.001 | .004 |
| Tobacco Use | -.034 | .427 | 1.0 |
| Social Advantage | .129 | .008 | .032 |
| Psychosocial Distress | -.023 | .597 | 1.0 |
| **Total Subcortical Gray Matter, $R^2 = .560$, $p < .001$** |  |  |  |
| Sex | -.244 | <.001 | .004 |
| Birthweight | .202 | <.001 | .004 |
| PMA at scan | .541 | <.001 | .004 |
| Tobacco Use | -.068 | .112 | .448 |
| Social Advantage | .152 | .002 | .008 |
| Psychosocial Distress | -.046 | .303 | 1.00 |
| **Total White Matter, $R^2 = .367$, $p < .001$** |  |  |  |
| Sex | -.290 | <.001 | .004 |
| Birthweight | .188 | <.001 | .004 |
| Tobacco Use | -.056 | .272 | 1.00 |
| PMA at scan | .187 | <.001 | .004 |
| Social Advantage | .284 | <.001 | .004 |
| Psychosocial Distress | -.024 | .651 | 1.00 |
| **Total Left Hippocampus, $R^2 = .218$, $p < .001$** |  |  |  |
| Sex | .162 | .003 | .012 |
| Birthweight | .115 | .048 | .192 |
| PMA at scan | .293 | <.001 | .004 |
| Tobacco Use | -.026 | .643 | 1.00 |
| Social Advantage | .170 | .009 | .036 |
| Psychosocial Distress | .016 | .784 | 1.00 |
| **Total Right Hippocampus, $R^2 = .219$, $p < .001$** |  |  |  |
| Sex | -.142 | .010 | .040 |
| Birthweight | .151 | .010 | .040 |
| PMA at scan | .262 | <.001 | .004 |
| Tobacco Use | -.058 | .312 | 1.00 |
| Social Advantage | .170 | .009 | .036 |
| Psychosocial Distress | .012 | .837 | 1.00 |
| **Total Left Amygdala, $R^2 = .411$, $p < .001$** |  |  |  |
| Sex | -.297 | <.001 | .004 |
| Birthweight | .130 | .010 | .040 |
| PMA at scan | .370 | <.001 | .004 |
| Tobacco Use | -.095 | .054 | .216 |
| Social Advantage | .198 | <.001 | .004 |
| Psychosocial Distress | .007 | .893 | 1.00 |
| **Total Right Amygdala, $R^2 = .416$, $p < .001$** |  |  |  |
| Sex | -.282 | <.001 | .004 |
| Birthweight | .175 | .001 | .004 |
| PMA at scan | .374 | <.001 | .004 |
| Tobacco Use | -.052 | .292 | 1.00 |
| Social Advantage | .195 | .001 | .004 |
| Psychosocial Distress | .005 | .928 | 1.00 |
| **Total Cortical Surface Area, $R^2 = .539$, $p < .001$** |  |  |  |
| Sex | -.253 | <.001 | .001 |
| Birthweight | .202 | <.001 | .001 |
| PMA at scan | .490 | <.001 | .001 |
| Tobacco Use | .043 | .329 | .392 |
| Social Advantage | .219 | <.001 | .001 |
| Psychosocial Distress | -.020 | .695 | .695 |

*Note.* Results for all steps of hierarchical regression models are shown in full in Supplementary Material, Table S3.
<sup>a</sup> Q-values represent p-values after correction for multiple comparisons using Benjamini-Hochberg False Discovery Rate procedure.

**Figure 1.**
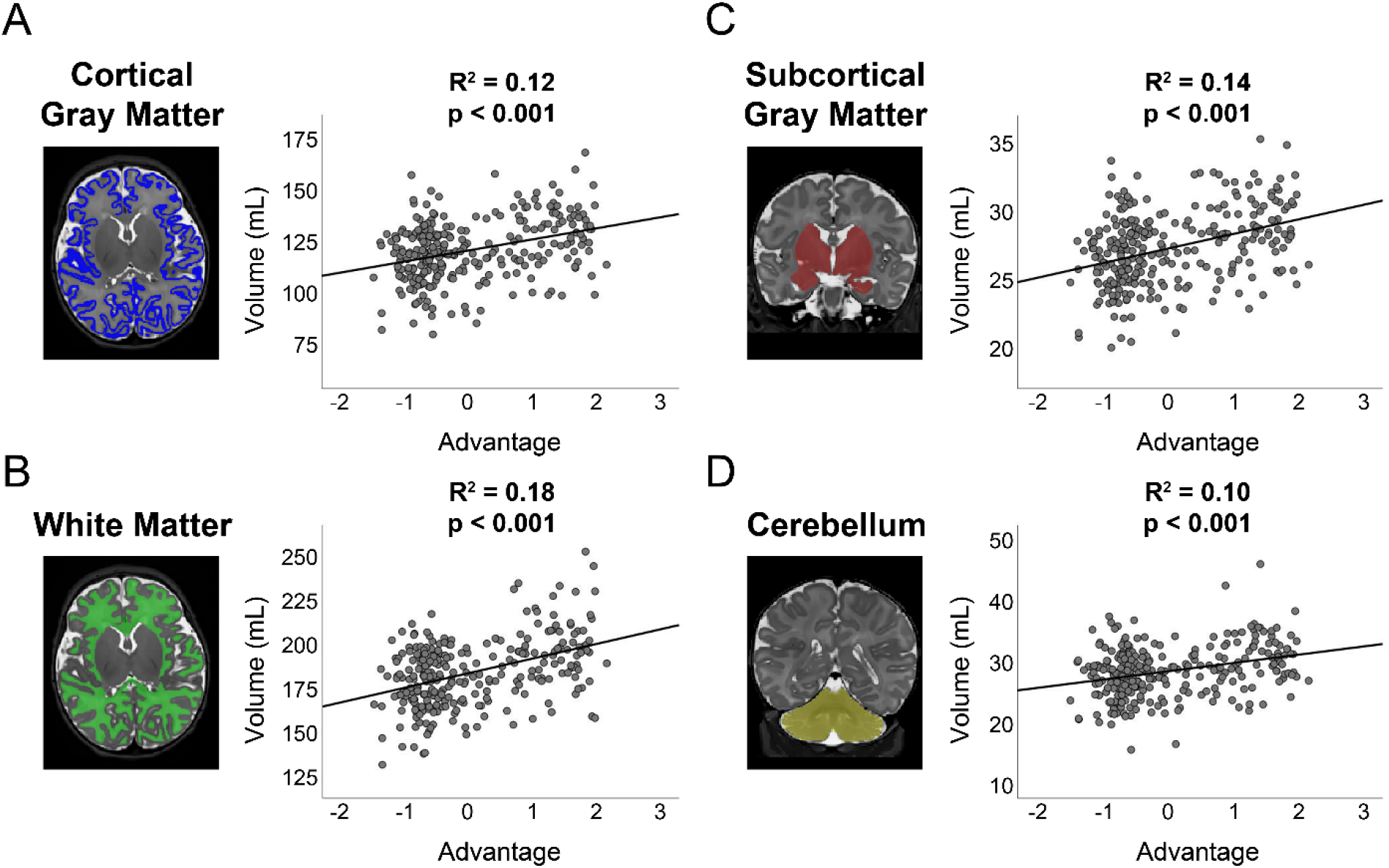
Plots demonstrating correlation between total brain volume and Maternal Social Advantage factor for A) cortical gray matter, B) white matter, C) subcortical gray matter, and D) cerebellum. Correlation and p values included for line of best fit included. Automated volumetric segmentation for each tissue type overlaid on T2-weighted image for representative infant also included.

#### Hippocampus and Amygdala

In Step 1, the following co-variates were related to smaller right and left hippocampi and amygdalae volumes: female sex, lower birthweight, and younger PMA at scan (Table S3). Tobacco use related to reduced amygdala volumes bilaterally (Table S3). In Step 2, greater social disadvantage was related to reduced volumes for subcortical regions of interest and accounted for an additional 2.0-3.1% of the variance (Table 2, Figure 2). The contribution of Psychosocial Stress was not significant (Table 2). After standardizing hippocampal and amygdalae volumes using TBV, there were no significant relationships with any co-variates or ELA constructs (Table S3).

**Figure 2.**
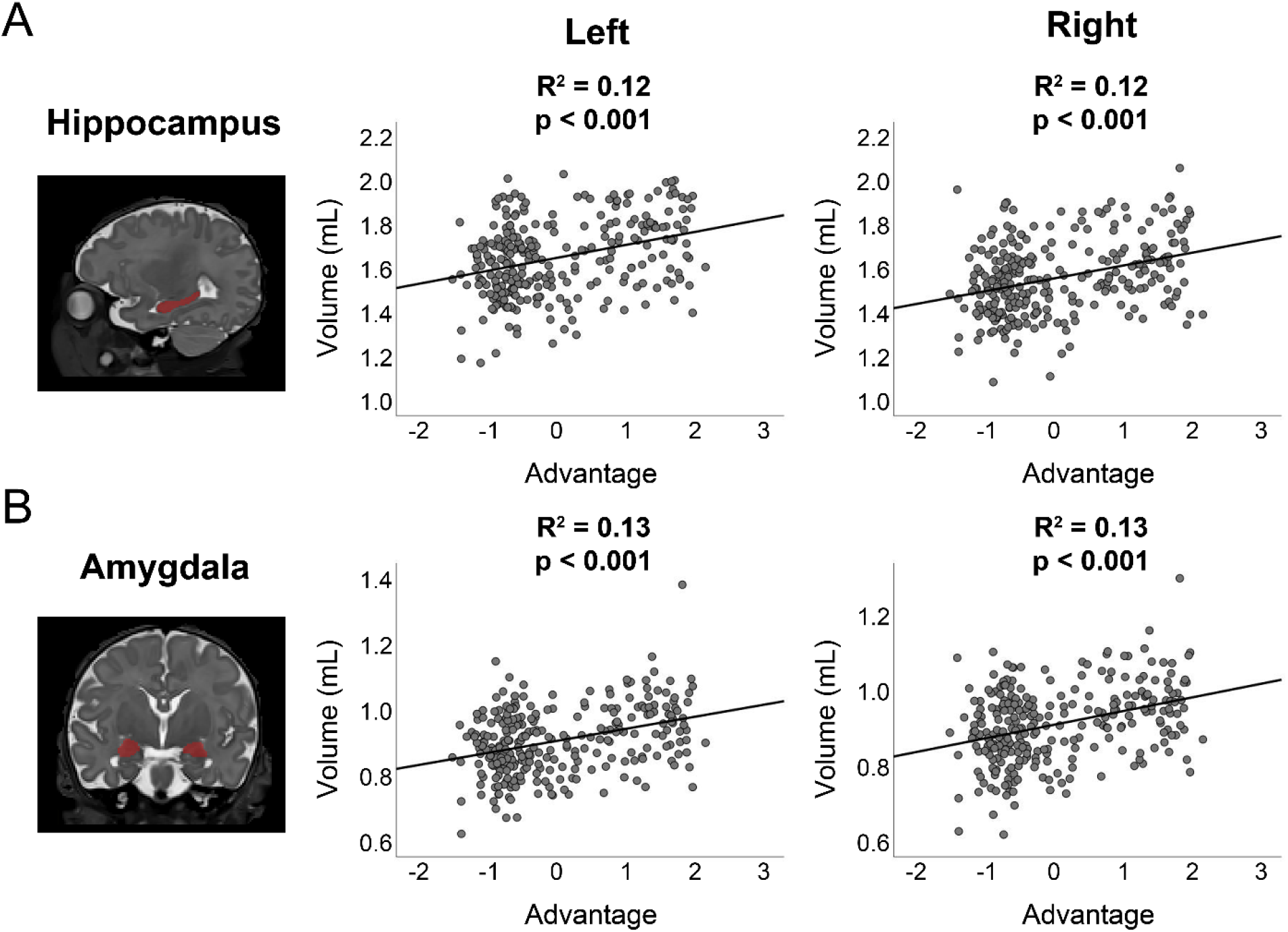
Plots demonstrating correlation between regional brain volume and Maternal Social Advantage factor for left and right A) hippocampus and B) amygdala. Correlation and p values included for line of best fit included. Automated volumetric segmentation for each structure overlaid on T2-weighted image for representative infant also included. Note similar results across hemispheres.

#### Cortical Folding

In Step 1, the following covariates related to diminished cortical folding: female sex, smaller birthweight, and younger PMA at scan (Table S3). In Step 2, higher social disadvantage related to reduced cortical folding and accounted for an additional 4.1% of the variance (Table 2, Figure 3). Tobacco use and Psychosocial Stress were not significant predictors.

**Figure 3.**
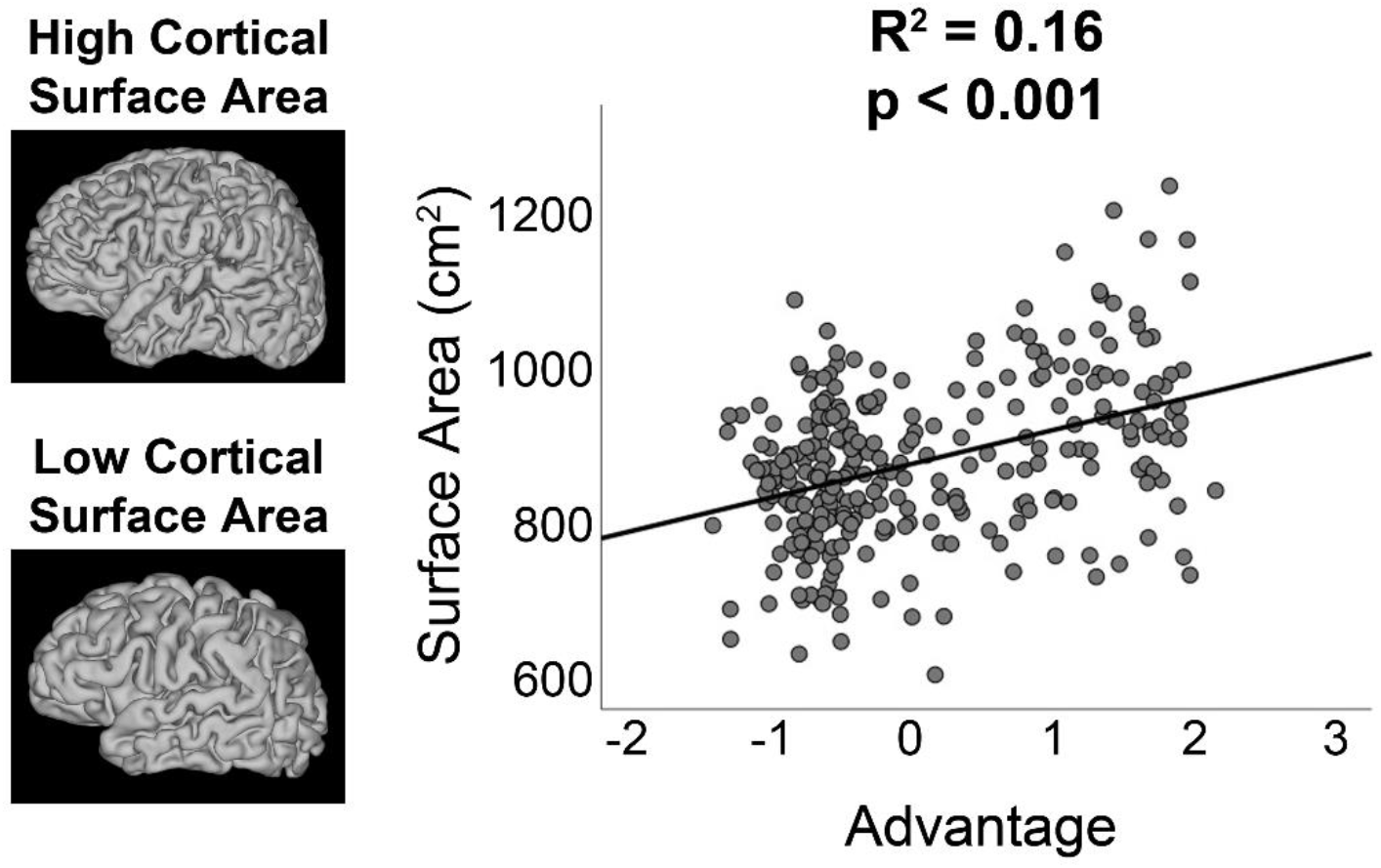
Plot demonstrating correlation between global cortical surface area (i.e., folding) and Maternal Social Advantage factor. Cortical surfaces for representative infants with high vs low cortical folding included for reference.

## Discussion

This is one of the largest investigations of the fetal origins of health and disease beginning in the first trimester of gestation with focus on disentangling the effects of prenatal exposure to poverty and maternal psychosocial stressors on brain morphometry at birth. Study findings suggest that in this cohort of healthy, term-born neonates, prenatal exposure to social disadvantage had a global adverse effect across all brain tissue types, including reduced cortical and subcortical gray and white matter, as well as decreased cortical folding in the first weeks of life. After accounting for global differences in brain volume, regionally-specific effects on the hippocampus and amygdala were limited. Potentially because this cohort was enriched for psychosocial adversity, exposure to higher levels of cumulative social disadvantage *in utero* appeared to play a greater role in brain structural development than maternal psychosocial distress.

We provide evidence for the global, adverse impact of prenatal exposure to socioeconomic disadvantage on brain structural growth and development at birth. Current findings are consistent with two cross-sectional studies showing that lower family I/R was related to reduced total cortical and subcortical gray matter in infants at five weeks^31^ and five months of age.^15^ Our findings are also consistent with work demonstrating regional reductions in cortical folding among older children from low socioeconomic backgrounds.^16^ Importantly, we show that the effects of poverty on brain volumes begin *in utero* and are evident on structural MRI in the first weeks of life. Among the different tissue types examined, social disadvantage was most strongly associated with reduced white matter volume, explaining 7% of the variance. This finding highlights the timing of prenatal exposure to poverty and the relative vulnerability of white matter, as myelination occurs rapidly beginning *in utero* at 28-29 weeks of gestation.^52,53^ During fetal development, oligodendrocyte progenitor cells and subplate neurons are selectively vulnerable to oxidative stress, and the deleterious impact of exposures may have cascading effects on pruning and/or crossing fibers and subsequent white matter volume at birth.^23,54^

Although social disadvantage *in utero* was associated with global reductions in brain volume and cortical folding at birth, the amygdala and hippocampus were not preferentially affected. Prenatal exposure to social disadvantage was related to reduced amygdala and hippocampus volume, but the association did not persist after adjustment for individual differences in total brain volume, which were also related to social disadvantage (Table S4). Previous work has shown the structural development of subcortical brain regions may be disproportionately susceptible to early exposure to poverty.^15,31^ Differences in study findings may be attributed to the fact that prior work has relied on single measures of adversity, assessed brain development at later timepoints, and/or included samples of higher socioeconomic status.^15,28,31,55^ Thus, we interpret the current study finding as evidence of a more widespread neuropathological disruption to global brain growth and development in the setting of exposure to significant, multifactorial socioeconomic disadvantage *in utero*.

This study addresses the independent contributions of maternal social disadvantage and psychosocial distress during pregnancy on offspring brain morphometry at birth.^56^ Consistent with other findings,^57^ social disadvantage was modestly associated with psychological distress during pregnancy. However, prenatal exposure to social disadvantage was more strongly associated with brain volumes and cortical folding than maternal psychosocial stress, which was not significant. Current results could reflect the fact that participants were oversampled for mothers experiencing socioeconomic disadvantage. We also assessed multiple aspects of social adversity, which when examined together, are likely more impactful.^58,59^ Although the precise mechanism remains unclear, post-natal ELA studies posit that chronic deprivation of resources overstimulates the hypothalamic-pituitary-adrenal (HPA) axis and the immune system, leading to impaired brain-behavior outcomes.^13,60^ Fetal sensitivity to glucocorticoids during development is a leading hypothesis to explain the deleterious regional effects of prenatal ELA on the hippocampi, amygdalae, and prefrontal cortex.^35,61,62,63,64^ Alternatively, mothers experiencing multiple social hardships demonstrate altered cortisol production and systemic inflammation and often live in areas with higher levels of environmental pollutants, which together may differentially affect the maternal and developing fetal HPA axes^9,13,65,66^ Furthermore, maternal immune activation may contribute to globally impaired brain development *in utero* via mechanisms including increased synaptic pruning, altered neurotransmitter profiles, impaired placental delivery of neurotrophic factors, and placental epigenetic programming.^67,68^ Future directions to elucidate causal mechanisms of neurodevelopmental and socioemotional impairments include examining maternal inflammatory cytokines and cortisol,^55,61^ as well as linking prenatal ELA and brain morphometry findings to childhood outcomes.^69,70^

Unique strengths of the current study include a prospective, longitudinal design from the first trimester of pregnancy, a cohort enriched for social disadvantage, and a comprehensive multidimensional assessment of maternal social disadvantage and related psychosocial stressors. However, our findings should be interpreted in light of study limitations. First, we assessed maternal depression with the EPDS, and although this questionnaire is a validated measure of perinatal depression, the lack of a semi-structured interview may have led to underreporting of maternal depression. Second, this study did not assess other environmental exposures such as lead and air pollution, which may be linked with both poverty and aberrant brain development in offspring. Third, as described in Luby et al.,^37^ we did not investigate the role of race in this analysis due to the co-linearity between race and social disadvantage. This sample reflects the clear link between racial disparities and social disadvantage in the United States and provides justification for including a measure of racial discrimination.

In this study, we examined the independent roles of maternal social advantage and psychosocial distress during pregnancy. Key findings highlighted the adverse, global consequences of maternal social disadvantage on neonatal brain volumetric and folding measures, while the effects of maternal psychosocial distress in the setting of disadvantage were less prominent. Most importantly, these results highlight that the deleterious effects of poverty begin *in utero* and are evident in the first weeks of life. These findings may inform future randomized-controlled trials of poverty reduction and family-based interventions to address the psychosocial and material needs of expectant parents and prevent adverse neonatal brain outcomes at birth.^71^

## Data Availability

All data produced in the present work are contained in the manuscript.

