## Supplement for "Prenatal Exposure to Early Life Adversity and Neonatal Brain Volumes at Birth"

### SUPPLEMENTAL TABLES

**Table S1.** Identification of potential covariates of interest associated with infant sex (N=280)

| **Variable, mean (SD)** | **Males (n=149)** | **Females (n=131)** | ***t*** | ***p*** |
|---|---|---|---|---|
| Birthweight (g) | 3316 (470) | 3191 (500) | 2.159 | **.032** |
| PMA at Scan (weeks) | 41.74 (1.27) | 41.58 (1.30) | 1.074 | .284 |
| Social Advantage | .088 (1.014) | -.002 (.917) | .784 | .434 |
| Psych | -.169 (.875) | .049 (.891) | -1.135 | .257 |

**Table S2.** Identification of covariates of interest associated with neonatal volumetric MRI measures at birth (N=280).

| | **Maternal BMI*** | | **MMR Score** | | **Maternal Cannabis Use** | | **Maternal Tobacco Use** | | **Infant Birthweight** | | **Infant PMA at scan** | | **Infant Sex** | |
|---|---|---|---|---|---|---|---|---|---|---|---|---|---|---|
| **Volume** | ***r*** | ***p*** | ***r*** | ***p*** | ***F*** | ***p*** | ***F*** | ***p*** | ***r*** | ***p*** | ***r*** | ***p*** | ***t*** | ***p*** |
| TBV | -.067 | .325 | -.036 | .548 | .495 | .610 | 7.01 | **.001** | .432 | **<.001** | .519 | **<.001** | 6.33 | **<.001** |
| Total cGM | -.055 | .422 | -.057 | .343 | .262 | .770 | 5.07 | **.007** | .440 | **<.001** | .618 | **<.001** | 5.48 | **<.001** |
| Total subcortical GM | -.077 | .261 | -.047 | .436 | 1.08 | .341 | 7.12 | **.001** | .410 | **<.001** | .618 | **<.001** | 5.53 | **<.001** |
| Total WM | -.075 | .271 | -.038 | .531 | .644 | .526 | 7.47 | **.001** | .373 | **<.001** | .307 | **<.001** | 6.19 | **<.001** |
| Total Cerebellum | -.081 | .235 | -.039 | .571 | .755 | .471 | 4.30 | **.015** | .372 | **<.001** | .678 | **<.001** | 5.24 | **<.001** |
| Total Cortical SA | -.067 | .325 | -.036 | .548 | .424 | .655 | 2.76 | **.065** | .432 | **<.001** | .519 | **<.001** | 5.56 | **<.001** |

MMR = Maternal Medical Risk, PMA = postmenstrual age, TBV = total brain volume, cGM = cortical gray matter, GM = gray matter, WM = white matter, SA = surface area. ***** Maternal pre-pregnancy BMI data available for N=215.

**Table S3.** Full Results of the Hierarchical Regression Models Linking Maternal Social Advantage and Psychosocial Stress with Structural MRI Measures at Birth (*n*=280)

| | **Step 1** | | **Step 2** | | | **Change Statistics** | |
|---|---|---|---|---|---|---|---|
| **VOLUMES** | **β** | **p** | **β** | **p** | **FDR-adjusted q** | **$\Delta R^2$** | **p** |
| **Cortical Gray Matter** | $R^2$ = .543, p < .001 | | $R^2$ = .559, p < .001 | | | | |
| Sex | -.231 | <.001 | -.235 | <.001 | .004 | | |
| Birthweight | .286 | <.001 | .247 | <.001 | .004 | | |
| PMA at scan | .540 | <.001 | .522 | <.001 | .004 | | |
| Tobacco Use | -.072 | .082 | -.034 | .427 | 1.00 | | |
| Social Advantage | | | .129 | .008 | .032 | .016 | .008 |
| Psychosocial Distress | | | -0.023 | 0.597 | 1.00 | | |
| **Subcortical Gray Matter** | $R^2$ = .535, p < .001 | | $R^2$ = .560, p < .001 | | | | |
| Sex | -.232 | <.001 | -.236 | <.001 | .004 | | |
| Birthweight | .250 | <.001 | .201 | <.001 | .004 | | |
| PMA at scan | .545 | <.001 | .523 | <.001 | .004 | | |
| Tobacco Use | -.116 | .006 | -.068 | .112 | .448 | | |
| Social Advantage | | | .152 | .002 | .008 | .025 | <.001 |
| Psychosocial Distress | | | -.046 | .303 | 1.00 | | |
| **White Matter** | $R^2$ = .298, p < .001 | | $R^2$ = .367, p < .001 | | | | |
| Sex | -.281 | < .001 | -.290 | < .001 | .004 | | |
| Birthweight | .272 | < .001 | .188 | < .001 | .004 | | |
| PMA at scan | .226 | < .001 | .187 | < .001 | .004 | | |
| Tobacco Use | -.136 | .009 | -.056 | .272 | 1.0 | | |

| | | | | | | | |
|---|---|---|---|---|---|---|---|
| Social Advantage | | | .284 | < .001 | .004 | .069 | < .001 |
| Psychosocial Distress | | | -.024 | .651 | 1.0 | | |
| **Cerebellum** | $R^2$ = .569, p < .001 | | $R^2$ = .587, p < .001 | | | | |
| Sex | -.228 | < .001 | -.229 | < .001 | .004 | | |
| Birthweight | .206 | < .001 | .171 | < .001 | .004 | | |
| PMA at scan | .618 | < .001 | .604 | < .001 | .004 | | |
| Tobacco Use | -.043 | .288 | -.006 | .893 | 1.0 | | |
| Social Advantage | | | .093 | .049 | .196 | .017 | .004 |
| Psychosocial Distress | | | -.078 | .072 | .288 | | |
| | **Step 1** | | **Step 2** | | **FDR-adjusted** | **Change Statistics** | |
| **REGIONS OF INTEREST** | **β** | **p** | **β** | **p** | **q** | **$\Delta R^2$** | **p** |
| **Left Hippocampus** | $R^2$ = .197, p < .001 | | $R^2$ =.218, p < .001 | | | | |
| Sex | -.156 | .005 | .162 | .003 | .012 | | |
| Birthweight | .162 | .004 | .115 | .048 | .192 | | |
| PMA at scan | .316 | < .001 | .293 | < .001 | .004 | | |
| Tobacco Use | -.069 | .209 | -.026 | .643 | 1.00 | | |
| Social Advantage | | | .170 | .009 | .036 | .021 | .025 |
| Psychosocial Distress | | | .016 | .784 | 1.00 | | |
| **Right Hippocampus** | $R^2$ =.197, p < .001 | | $R^2$ = .219, p < .001 | | | | |
| Sex | -.136 | .014 | -.142 | .010 | .040 | | |

| | | | | | | | |
|---|---|---|---|---|---|---|---|
| Birthweight | .198 | <.001 | .151 | .010 | .040 | | |
| PMA at scan | .285 | <.001 | .262 | <.001 | .004 | | |
| Tobacco Use | -.101 | .067 | -.058 | .312 | 1.00 | | |
| Social Advantage | | | .170 | .009 | .036 | .022 | .023 |
| Psychosocial Distress | | | .012 | .837 | 1.00 | | |
| **Left Amygdala** | $R^2$ = .381, p < .001 | | $R^2$= .411, p < .001 | | | | |
| Sex | -.290 | < .001 | -.297 | <.001 | .004 | | |
| Birthweight | .186 | <.001 | .130 | .010 | .040 | | |
| PMA at scan | .397 | <.001 | .370 | <.001 | .004 | | |
| Tobacco Use | -.147 | .003 | -.095 | .054 | .216 | | |
| Social Advantage | | | .198 | <.001 | .004 | .030 | .001 |
| Psychosocial Distress | | | .007 | .893 | 1.00 | | |
| **Right Amygdala** | $R^2$ = .387, p < .001 | | $R^2$ = .416, p < .001 | | | | |
| Sex | -.275 | <.001 | -.282 | <.001 | .004 | | |
| Birthweight | .230 | <.001 | .175 | .001 | .004 | | |
| PMA at scan | .401 | <.001 | .374 | <.001 | .004 | | |
| Tobacco Use | -.103 | .033 | -.052 | .292 | 1.00 | | |
| Social Advantage | | | .195 | .001 | .004 | .030 | .001 |
| Psychosocial Distress | | | .005 | .928 | 1.00 | | |
| **Relative Left Hippocampus [a]** | $R^2$ .010, p = .405 | | $R^2$ = .015, p = .543 | | | | |

| | | | | | | | |
|---|---|---|---|---|---|---|---|
| Sex | .074 | .224 | .072 | .239 | .096 | | |
| PMA at scan | -.055 | .358 | -.049 | .430 | 1.00 | | |
| Tobacco Use | .028 | .642 | .012 | .851 | 1.00 | | |
| Social Advantage | | | -.024 | .731 | 1.00 | .004 | .568 |
| Psychosocial Distress | | | .053 | .427 | 1.00 | | |
| **Relative Right Hippocampus** [a] | $R^2$ = .024, p = .085 | | $R^2$ = .027, p = .189 | | | | |
| Sex | .112 | .064 | .110 | .069 | .276 | | |
| PMA at scan | -.100 | .096 | -.094 | .123 | .482 | | |
| Tobacco Use | -.010 | .069 | -.018 | .793 | 1.00 | | |
| Social Advantage | | | -.018 | .793 | 1.00 | .003 | .649 |
| Psychosocial Distress | | | .047 | .472 | 1.00 | | |
| **Relative Left Amygdala** [a] | $R^2$ = .012, p = .344 | | $R^2$ = .022, p = .297 | | | | |
| Sex | -.039 | .520 | -.041 | .494 | 1.0 | | |
| PMA at scan | -.076 | .206 | -.062 | .312 | 1.0 | | |
| Tobacco Use | -.069 | .254 | -.098 | .124 | .496 | | |
| Social Advantage | | | -.059 | .398 | 1.0 | .010 | .252 |
| Psychosocial Distress | | | .065 | .322 | 1.0 | | |
| **Relative Right Amygdala** [a] | $R^2$ = .004, p = .611 | | $R^2$ - .011, p = .555 | | | | |
| Sex | -.013 | .826 | -.016 | .791 | 1.00 | | |
| PMA at scan | -.059 | .329 | -.049 | .427 | 1.00 | | |

| | | | | | | | |
|---|---|---|---|---|---|---|---|
| Tobacco Use | .000 | .997 | -.023 | .719 | 1.00 | | |
| Social Advantage | | | -.038 | .591 | 1.00 | .008 | .341 |
| Psychosocial Distress | | | .070 | .294 | 1.00 | | |
| | **Step 1** | | **Step 2** | | **FDR-adjusted** | **Change Statistics** | |
| **SURFACE AREA** | **β** | **p** | **β** | **p** | **q** | **$\Delta R^2$** | **p** |
| **Total Surface Area** | $R^2$ = .498, p < .001 | | $R^2$ = .539, p < .001 | | | | |
| Sex | -.246 | <.001 | -.253 | <.001 | .001 | | |
| Birthweight | .267 | <.001 | .202 | <.001 | .001 | | |
| PMA at scan | .520 | <.001 | .490 | <.001 | .001 | | |
| Tobacco Use | -.019 | .661 | .043 | .329 | .392 | | |
| Social Advantage | | | .219 | <.001 | .001 | 0.041 | <.001 |
| Psychosocial Distress | | | -.20 | .658 | .695 | | |

[a] Birthweight was not included as an independent variable for relative region of interest volumes adjusted for total brain volume to avoid overfitting the regression models. Relative region of interest volumes were computed as the volume of the region divided by total brain volume.

**Table S4.** Hierarchical Regression Model Linking Maternal Social Advantage and Psychosocial Stress with Total Brain Volumes (TBV) at Birth (*n*=280)

| | **Step 1** | | **Step 2** | | **Change Statistics** | |
|---|---|---|---|---|---|---|
| **Total Brain Volume** | **β** | **p** | **β** | **p** | **Δ$R^2$** | **p** |
| | $R^2$ = .470, p < .001 | | $R^2$ = .512, p < .001 | | | |
| Sex | -.275 | < .001 | -.281 | < .001 | | |
| Birthweight | .290 | < .001 | .405 | < .001 | | |
| PMA at scan | .435 | < .001 | .225 | < .001 | | |
| Tobacco Use | -.109 | .015 | -.046 | .302 | | |
| Social Advantage | | | .214 | < .001 | .042 | < .001 |
| Psychosocial Distress | | | -.034 | .474 | | |

**SUPPLEMENTAL FIGURES**

**Figure S1.** Participant flow from study enrollment to inclusion in current analysis.

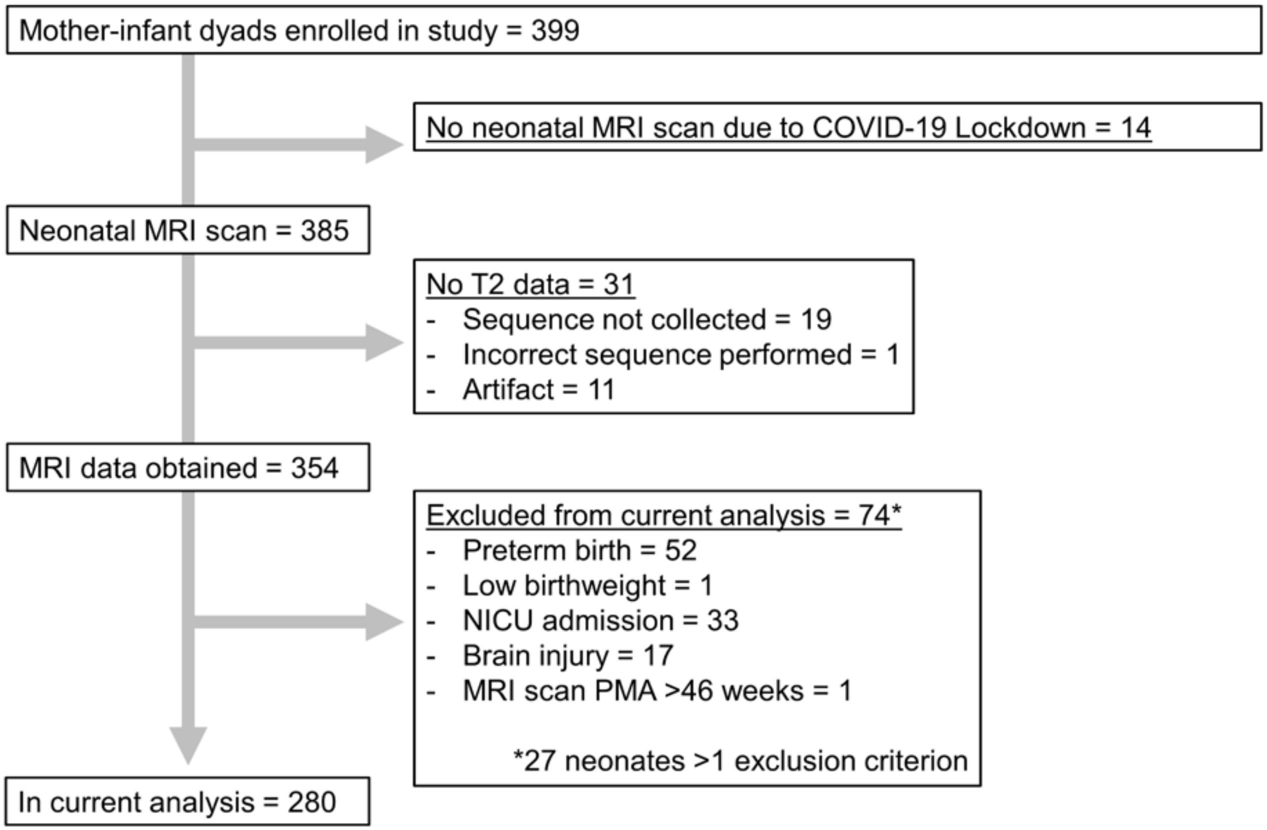
